# Smoking methylation marks for prediction of urothelial cancer risk

**DOI:** 10.1101/2021.03.16.21253681

**Authors:** Chenglong Yu, Kristina M. Jordahl, Julie K. Bassett, Jihoon Eric Joo, Ee Ming Wong, Maree Brinkman, Daniel Schmidt, Damien Bolton, Enes Makalic, Theodore M. Brasky, Aladdin H. Shadyab, Lesley Tinker, Anthony Longano, John L. Hopper, Dallas R. English, Roger L. Milne, Parveen Bhatti, Melissa C. Southey, Graham G. Giles, Pierre-Antoine Dugué

## Abstract

**Background:** Self-reported information may not accurately capture smoking exposure. We aimed to evaluate whether smoking-associated DNA methylation markers improve urothelial cell carcinoma (UCC) risk prediction.

**Methods:** Conditional logistic regression was used to assess associations between blood-based methylation and UCC risk using two matched case-control samples, *N*=404 pairs from the Melbourne Collaborative Cohort Study (MCCS) and *N*=440 pairs from the Women’s Health Initiative (WHI) cohort, respectively. Results were pooled using fixed-effects meta-analysis. We developed methylation-based predictors of UCC and evaluated their prediction accuracy on two replication datasets using the area under the curve (AUC).

**Results:** The meta-analysis identified associations (P<4.7×10^−5^) for 29 of 1,061 smoking-associated methylation sites, but these were substantially attenuated after adjustment for self-reported smoking. Nominally significant associations (P<0.05) were found for 387 (36%) and 86 (8%) of smoking-associated markers without/with adjustment for self-reported smoking, respectively, with same direction of association as with smoking for 387 (100%) and 79 (92%) markers. A Lasso-based predictor was associated with UCC risk in one replication dataset in MCCS (*N*=134, odds ratio per SD [OR]=1.37, 95%CI=1.00-1.90) after confounder adjustment; AUC=0.66, compared with AUC=0.64 without methylation information. Limited evidence of replication was found in the second testing dataset in WHI (*N*=440, OR=1.09, 95%CI=0.91-1.30).

**Conclusions:** Combination of smoking-associated methylation marks may provide some improvement to UCC risk prediction. Our findings need further evaluation using larger datasets.

**Impact:** DNA methylation may be associated with UCC risk beyond traditional smoking assessment and could contribute to some improvements in stratification of UCC risk in the general population.

## Introduction

Urothelial cell carcinoma (UCC) is a type of malignancy arising from the urothelium. While UCC accounts for more than 90% of urinary bladder cancers (1), some can also be found in the proximal urethra, the transitional epithelium of the renal pelvis, and the ureter (2). According to Global Cancer Statistics 2018, bladder cancer is the tenth most common cancer worldwide, with an estimated 549,000 new cases and 200,000 deaths (3). Cigarette smoking has been established as a strong risk factor for UCC with approximately half of newly diagnosed patients reporting a history of smoking (4, 5). Many studies (6-9) have investigated the association between smoking and risk of UCC, and a meta-analysis of 89 observational studies reported an increased risk of bladder cancer for current smokers (odds ratio [OR] = 3.1, 95% confidence interval [CI] = 2.5-3.7) and former smokers (OR = 1.8, 95% CI = 1.5-2.1), compared with never smokers (10). However, information on smoking history used in most epidemiological studies, such as smoking status (never, former or current smoker) or pack-years, is typically collected via self-report and may be prone to substantial measurement error. The accuracy of self-reported information has also been questioned because of declining response rates and the increasing social stigmatisation of smoking (11). Furthermore, such information cannot reflect second-hand smoke exposure during childhood or adulthood. Therefore, such less accurate information would have potential impact on studies of disease association and risk prediction.

Serum or urinary cotinine (12) and blood DNA methylation (13-16) have been established as valid biomarkers of cigarette smoking exposure. Although cotinine and methylation markers showed similar accuracy in distinguishing current from never smokers, only methylation markers can distinguish former from never smokers with high accuracy (17). Therefore, DNA methylation markers measured in blood, which may also reflect different individuals’ responses to lifetime exposure, can be used to augment self-reported smoking data to help refine individual risk profiling of smoking-induced diseases (18-20).

Authors of several studies (21-23) have evaluated the association of genome-wide cytosine-guanine (CpG) methylation in blood DNA with risk of UCC. Jordahl et al. (23), for example, identified potential methylation-based markers of susceptibility to urothelial carcinoma of the bladder, using the Illumina Infinium HumanMethylation450 Bead Array (∼450,000 probes) on pre-diagnostic blood collected in the Women’s Health Initiative (WHI). They subsequently found that two previously identified smoking-associated CpG sites mediated the effect of smoking on bladder cancer risk (24). With the current study, we aimed to expand on previous research by identifying associations between smoking-associated DNA methylation and bladder cancer risk and by developing a predictor of UCC risk using smoking-associated DNA methylation measures.

## Materials and Methods

### Study participants

The Melbourne Collaborative Cohort Study (MCCS) is an Australian prospective cohort study of 41,513 people recruited between 1990 and 1994 in the Melbourne metropolitan area. All participants were of white European origin. DNA was extracted from pre-diagnostic peripheral blood taken at recruitment (1990-1994) or at a subsequent follow-up visit (2003-2007) in participants free of UCC. More details about the cohort, blood collection, DNA extraction and cancer ascertainment can be found elsewhere (22, 25). In this study, we utilized a case-control data set of urothelial cancer nested within the MCCS. Controls were matched to incident cases on age at blood draw, year of birth, sex, country of birth (Australia/New-Zealand/UK/other, Italy or Greece), sample collection period (baseline at recruitment or the follow-up visit) and sample type (peripheral blood mononuclear cells, dried blood spots or buffy coats) using incidence density sampling. To minimise batch effects, samples from each matched case-control pair were plated to adjacent wells on the same BeadChip, with plate, chip, and position assigned randomly. We excluded from the analysis sex-discrepant and failed samples for DNA methylation measures. Case-control pairs with any missing values for the confounders measured were also excluded. Overall, 404 case-control pairs were included in the present study.

For replication and meta-analysis, we included the study sample previously used by Jordahl et al. (23, 24), which comprises 440 cases diagnosed with urothelial carcinoma of the bladder and 440 cancer-free controls matched on year of enrollment, age at enrollment (±2 years), follow-up time greater than or equal to their matched case, trial component and DNA extraction method (5-Prime, phenol, Bioserve, or PurGene). This case-control study was nested within the WHI, which includes 161,808 postmenopausal women recruited from 1993 to 1998 across the United States (26).

The study was approved by the Cancer Council Victoria’s Human Research Ethics Committee, Melbourne, VIC, Australia, and the Institutional Review Board and Publications and Presentations Committee of WHI - Clinical Coordinating Center in the Fred Hutchinson Cancer Research Center, Seattle, WA, USA. All participants provided informed consent in accordance with the Declaration of Helsinki.

### Quality control and normalisation of methylation data

Quality control (QC) details for measures of genome-wide DNA methylation in the MCCS have been reported previously (22). Briefly, we removed probes with missing rate > 20% and probes on Y-chromosome, and ultimately retained 484,966 CpG sites with their beta values for each sample. Methylation M-values, calculated as log_2_(beta/(1-beta)), were used for analysis as these are thought to be more statistically valid for detection of differential methylation (27). In the replication data of WHI, similar data processing on DNA methylation were performed, e.g. QC on CpGs sites using probe missing rate (> 10%) and beadcount (<3) in at least 10% of samples, and M-value transformation, as described previously (23, 24).

### Association analysis of genome-wide DNA methylation

An epigenome-wide association study (EWAS) based on the 404 case-control pairs in MCCS was conducted, using conditional logistic regression to estimate OR and 95% CI of UCC risk per SD at each of the 484,966 CpG sites. A first model (Model 1) was adjusted for white blood cell composition (percentage of CD4 + T cells, CD8 + T cells, B cells, NK cells, monocytes and granulocytes, estimated using the Houseman algorithm [28]), and a second model (Model 2) was additionally adjusted for smoking status (current / former / never) and pack-years (log-transformed). As a sensitivity analysis, we evaluated a third model (Model 3) with additional adjustment for alcohol consumed in the previous week (in grams/day), body mass index (in kg/m^2^), height (in metres), educational level (pseudo-continuous score ranging from 1 for “primary school only” to 8 for “tertiary or higher university degree”), physical activity (categorised score based on time spent doing vigorous/less vigorous activities), socioeconomic status (deciles of the relative socioeconomic disadvantage of area of residence index) and diet quality (Alternative Healthy Eating Index 2010). We also stratified analyses by sex and clinical subtype (muscle invasive or non-muscle invasive) and tested heterogeneity of the associations using the likelihood ratio test, by comparing models with and without interaction terms for these variables. The Bonferroni correction was applied to account for multiple comparisons (P<0.05/484966 =1.03×10^−7^).

### Association analysis of smoking-associated DNA methylation

Among the 484,966 probes, we focused on 1,061 sites that were found to be strongly associated with a comprehensive smoking index in the MCCS (P<10^−7^) and also reported to be associated with smoking at this threshold P<10^−7^ in any of six large studies, as described in our previous publication (ref. 29; see Supplementary Table 1). For the replication study, we also used conditional logistic regression (Model 1 and Model 2) to estimate associations of the 1,061 smoking-associated DNA methylation measures with risk of UCC in the WHI. For the WHI study, Model 1 and Model 2 were additionally adjusted for race/ethnicity (Asian/Pacific Islander, Black/African American, Hispanic/Latino, non-Hispanic White, or other). The Bonferroni correction was applied to account for multiple comparisons (P<0.05/1061 =4.7×10^−5^).

### Meta-analysis of MCCS and WHI studies

A fixed-effects meta-analysis with inverse-variance weights was conducted to combine associations with UCC risk at the 1,061 smoking-associated CpGs from the analyses of MCCS and WHI, using the *metagen* function in the R package *meta* (30). The I-square statistic was used to assess heterogeneity across the two studies.

### Predictive models

A predictor of UCC risk was developed using the data of 270 case-control pairs from the MCCS cohort for which blood was collected at baseline (1990-1994) as the training set (discovery phase), and 134 case-control pairs for which blood was drawn at follow-up (2003-2007) as an independent testing set in the validation phase. We used penalised logistic regression models with UCC risk as the outcome and the M-values at the 1,061 smoking-associated CpGs as the independent variables, applied to the training set using the R package *glmnet* (31). Fivefold cross-validation was used, and the mixing parameter (alpha) was set to 1 to apply a Lasso (least absolute shrinkage and selection operator) penalty. The covariates used in Model 3 were forced in the penalised logistic models. Coefficients of the logistic Lasso model with the lambda value corresponding to the minimum mean cross-validated error were extracted and used as weights of the selected CpGs to construct a smoking methylation score (MS) for each participant. The smoking MS was then evaluated as a predictor of UCC risk in conditional logistic regression models (adjusted for covariates in Model 3 for MCCS data and in Model 2 for WHI data, respectively) in the validation sets.

Alternative ways to build methylation-based predictors of UCC risk were explored. We conducted univariate analyses using conditional logistic regression models to the training set to estimate ORs for the individual associations between DNA methylation and UCC risk at each of the 1,061 CpGs. The same covariates as those forced in the Lasso models were included as covariates. We considered three P-value cut-offs (0.05, 0.01 and 0.001) of individual associations at the 1,061 sites, and for each of them we calculated a smoking MS as a weighted average using as weights the logarithm of the OR for each selected CpG.

As a sensitivity analysis, we also used the logistic Lasso method (as described above) to develop a DNA methylation-based smoking predictor of UCC risk using all 404 MCCS case-control pairs. The external 440 case-control pairs from the WHI study were then used as an independent validation set to assess the proposed DNA methylation-based smoking predictors by using conditional logistic regression models (adjusted for covariates in Model 2).

The accuracy of the predictive models with the smoking MS as UCC risk predictor was assessed using area under the receiver operating characteristic curve (AUC) estimates with unconditional logistic regression models (Models A, B and C), using the R package *pROC* (32). Model A used white blood cell composition as independent variables. Model B used white blood cell composition, smoking status and pack-years (log-transformed) as independent variables. Race/ethnicity was also included in the two models for the WHI sample. Model C used white blood cell composition, smoking status, pack-years and other covariates (age, sex, country of birth, sample type, alcohol, BMI, height, educational level, physical activity, socioeconomic status and diet quality) as independent variables. The proposed methylation scores were then used as additional independent variables in the models to assess the prediction performance by AUC. The DeLong test (33) was used for comparing AUCs.

All methylation scores were rescaled to Z-scores for better comparability of their association with UCC risk. The flowchart of the statistical analysis pipelines and method details are shown in Figure 1.

**Figure 1.**
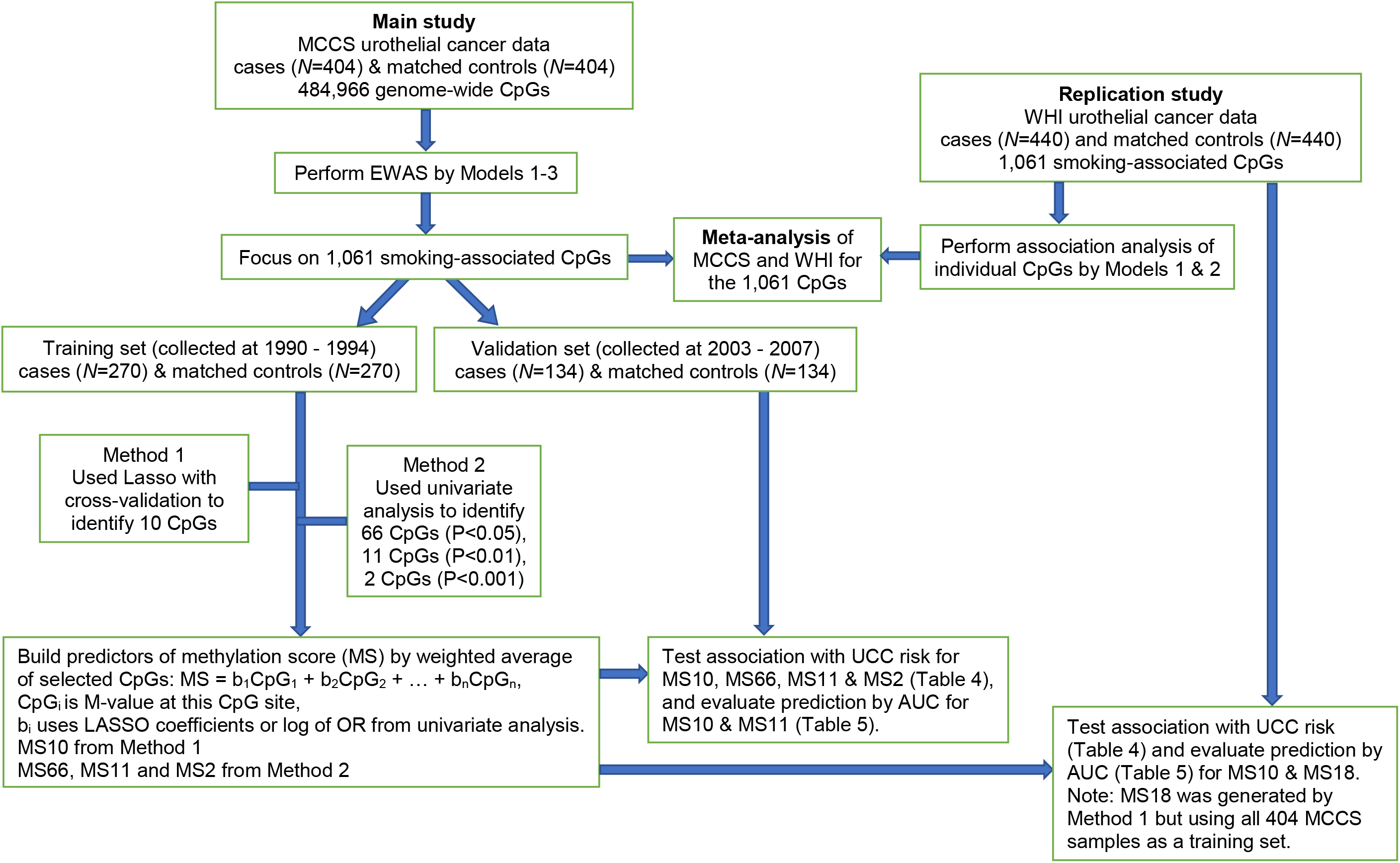
Flowchart of the study.

## Results

The distribution of sociodemographic, lifestyle, anthropometric and clinical characteristics of the participants in the MCCS is presented in Table 1. Controls were matched to cases on age at blood draw, sex, country of birth (Australia/New-Zealand/UK/other, Italy or Greece) and sample type (peripheral blood mononuclear cells, dried blood spots and buffy coats). The participants in the MCCS testing set were an average eight years older than in the training set. Compared with controls, cases were more frequently past and current smokers, and had greater smoking pack-years.

**Table 1:** Characteristics of the MCCS participants included in the analyses.

| Participant characteristics | Training set (1990-1994) |  | Testing set (2003-2007) |  |
| --- | --- | --- | --- | --- |
|  | UCC cases<br>(N=270) | Controls<br>(N=270) | UCC cases<br>(N=134) | Controls<br>(N=134) |
| Age at blood draw, median [IQR] | 63<br>[58-67] | 64<br>[58-67] | 72<br>[67-77] | 72<br>[67-77] |
| Sex: |  |  |  |  |
| male, N (%) | 207 (77%) | 207 (77%) | 101 (75%) | 101 (75%) |
| female, N (%) | 63 (23%) | 63 (23%) | 33 (25%) | 33 (25%) |
| Country of birth: |  |  |  |  |
| Australia/NZ/UK/other, N (%) | 168 (62%) | 166 (61%) | 104 (78%) | 104 (78%) |
| Italy, N (%) | 56 (21%) | 58 (21%) | 20 (15%) | 20 (15%) |
| Greece, N (%) | 46 (17%) | 46 (17%) | 10 (7%) | 10 (7%) |
| Blood sample type: |  |  |  |  |
| Dried blood spots, N (%) | 170 (63%) | 170 (63%) | 1 (1%) | 1 (1%) |
| Peripheral blood mononuclear cells, N (%) | 93 (34%) | 93 (34%) | 0 (0%) | 0 (0%) |
| Buffy coats, N (%) | 7 (3%) | 7 (3%) | 133 (99%) | 133 (99%) |
| Smoking: |  |  |  |  |
| current, N (%) | 51 (19%) | 41 (15%) | 22 (16%) | 13 (10%) |
| former, N (%) | 146 (54%) | 111 (41%) | 68 (51%) | 63 (47%) |
| never, N (%) | 73 (27%) | 118 (44%) | 44 (33%) | 58 (43%) |
| Smoking pack-years, median [IQR] | 18<br>[0-40.7] | 4.2<br>[0-29.6] | 11.4<br>[0-35.1] | 5.2<br>[0-19.8] |
| Height (cm), median [IQR] | 168<br>[162-173] | 168<br>[163-173] | 169<br>[162-176] | 170<br>[164-175] |
| Body mass index (kg/m2), median [IQR] | 27.5<br>[25.4-29.8] | 27.1<br>[24.8-29.5] | 27.3<br>[24.7-29.8] | 27.2<br>[24.5-29.5] |
| Alcohol (ethanol) consumption (g/day), median [IQR] | 4.5<br>[0-20.5] | 6.8<br>[0-17.7] | 9.2<br>[1.3-23.6] | 8.7<br>[0.6-23.4] |
| Diet quality: AHEI-2010, median [IQR] | 63.0<br>[55.0-70.9] | 64.5<br>[57.0-72.0] | 64.5<br>[55.0-70.5] | 63.0<br>[57.5-72.4] |
| Physical activity score, median [IQR] | 2<br>[1.3-2] | 2<br>[2-2] | 2<br>[2-3] | 2<br>[2-2.8] |
| Education score, median [IQR] | 4<br>[3-5] | 4<br>[3-6] | 4<br>[4-7] | 4<br>[4-8] |
| Socioeconomic status, SEIFA-10, median [IQR] | 5<br>[3-8] | 5<br>[3-8] | 6<br>[4-9] | 6<br>[3-9] |

Note: Physical activity score is a categorised score based on time spent doing vigorous/less vigorous activities. Educational score is a pseudo-continuous score ranging from 1 for “primary school only” to 8 for “tertiary or higher university degree”.

For the genome-wide probes tested on the 404 MCCS case-control pairs using Models 1-3, there was no significant association between DNA methylation and risk of UCC after Bonferroni correction (P<1.03×10^−7^). Nominally significant associations (P<0.05) were observed for 40,664 (∼8%), 32,137 (∼7%) and 31,319 (∼6%) of the 484,966 CpGs using Models 1-3, respectively.

Focusing on the 1,061 smoking-associated CpG sites that we previously identified (29), there was no significant association between DNA methylation and UCC risk in the MCCS after Bonferroni correction (P<4.7×10^−5^). Comparing to genome-wide results, there were more methylation markers associated with risk of UCC for the smoking-associated loci, e.g. 19 of the 25 CpGs most strongly with smoking had P<0.05 in Model 1 (Supplementary Table 1). Nominally significant associations (P<0.05) were observed for 206 (∼19%) and 93 (∼9%) of the 1,061 CpGs in Models 1 and 2, respectively (Supplementary Table 1), and the direction of the association was the same as for smoking for 205/206 (100%) and 88/93 (95%) CpG sites. Adjustment for a more comprehensive set of variables (Model 3) did not substantially change the associations (Table 2 and Supplementary Figure 1). Furthermore, the direction of association at 883 (83%, 662 negative and 221 positive, Model 1) and 766 (72%, 586 negative and 180 positive, Model 2) of the 1,061 CpGs was the same as for their association with smoking (Supplementary Table 1). The results for the 20 most significant associations are presented in Table 2; for all of these associations, the direction of association was the same as with smoking. The stratified results by UCC subtype and sex are shown in Supplementary Tables 2 and 3; we observed no evidence of significant UCC subtype or sex heterogeneity after Bonferroni correction for multiple testing (P<4.7×10^−5^).

**Table 2:**
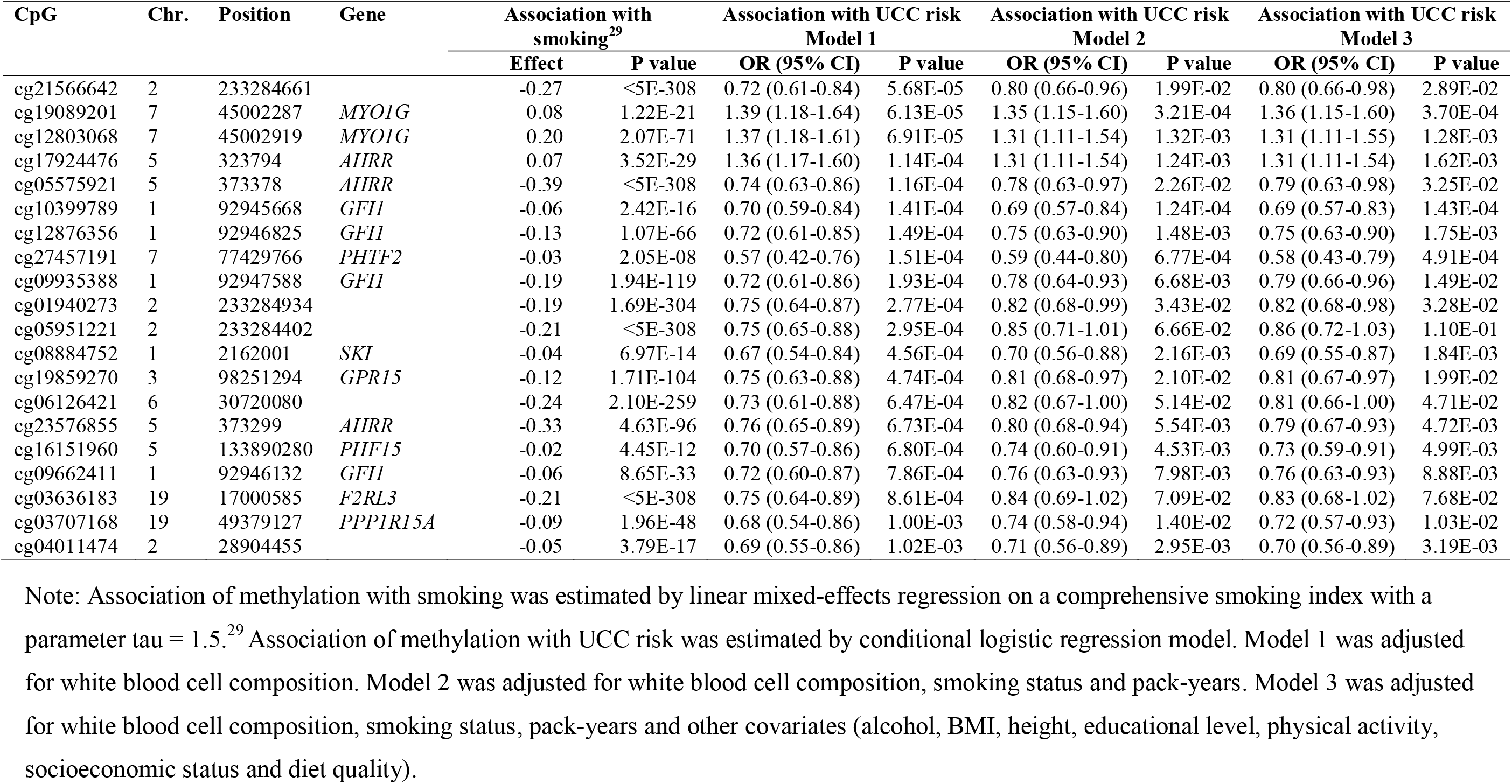
EWAS results of UCC risk in the 20 most significant associations (Model 1) of the 1,061 smoking-associated CpGs based on the 404 matched case-control pairs in the MCCS.

The replication study using WHI data identified nominally significant associations (P<0.05) for 229 (∼22%) and 47 (∼4%) of the 1,061 smoking-based CpGs in Models 1 and 2, respectively (Supplementary Tables 4 and 5). Among these associations, 51 CpGs (Model 1) and 3 CpGs (Model 2) were also nominally significant and in the same direction as in the MCCS data.

The meta-analysis of MCCS and WHI results identified nominally significant associations for 387 (∼36%) and 86 (∼8%) CpG sites in Models 1 and 2, respectively (Supplementary Tables 4 and 5), and the direction of the association was the same as the association with smoking for 387/387 (100%) and 79/86 (92%) of the CpGs. There were 29 significant associations in Model 1 after Bonferroni correction (P<4.7×10^−5^), and among these associations, 9 CpGs overlapping the *AHRR, GPR15, F2RL3, PRSS23* and *GFI1* genes were genome-wide significant (P<1.03×10^−7^). The associations were nevertheless substantially attenuated (all P>4.7×10^−5^) after adjusting for self-reported smoking variables (Model 2). For the majority of the 1,061 CpGs, there was little heterogeneity between MCCS and WHI results (81% and 83% of the CpGs had I^2^ < 0.5 in Models 1 and 2, respectively, see Supplementary Tables 4 and 5). The 20 strongest associations in the meta-analyses of Model 1 and Model 2 are shown in Table 3.

**Table 3:**
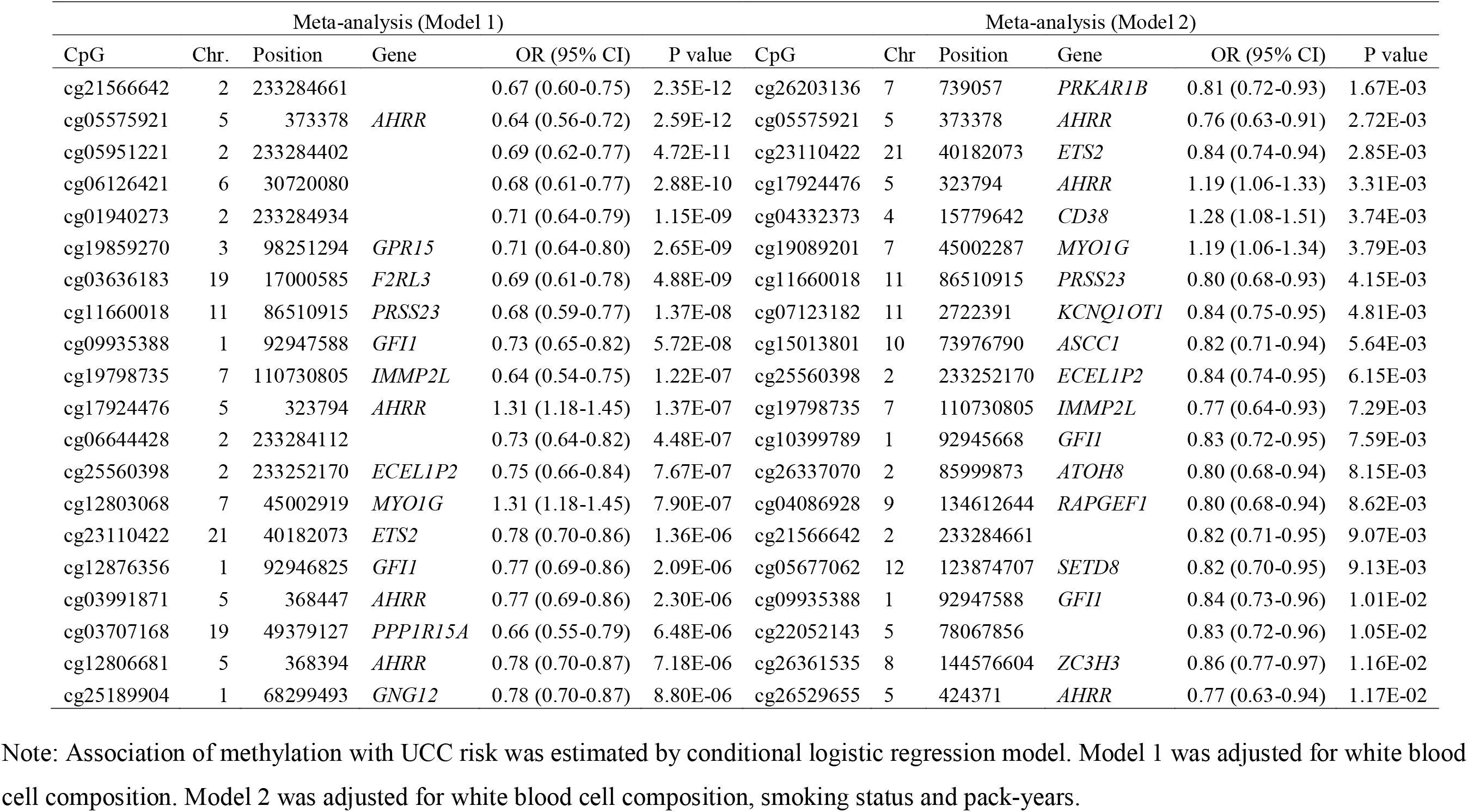
Meta-analysis results for MCCS and WHI in the 20 most significant associations between DNA methylation at 1,061 smoking-associated CpGs and risk of UCC by Model 1 and Model 2, respectively.

The logistic Lasso regression of UCC risk on the 1,061 smoking-based CpGs using the 270 MCCS baseline case-control pairs selected ten CpGs (MS10): cg01324550 (*LOC404266*), cg02743070 (*ZMIZ1*), cg07058086 (*KIF13B*), cg10399789 (*GFI1*), cg16622061 (chr16: 86888736), cg17924476 (*AHRR*), cg18979623 (*ZBTB46*), cg19089201 (*MYO1G*), cg23110422 (*ETS2*) and cg24139443 (chr17: 74131549) (Supplementary Table 6). The associations with risk of UCC for the 1,061 smoking-associated methylation sites on the training data are shown in Supplementary Table 6. The derived methylation scores based on associations at P<0.05, P<0.01 and P<0.001 included 66 (MS66), 11 (MS11) and 2 (MS2) CpGs, respectively. The associations of these four predictors with UCC risk in the MCCS validation dataset (*N*=134 cases, Model 3) are presented in Table 4. MS10 and MS11 had five overlapping CpGs (cg07058086, cg10399789, cg17924476, cg19089201 and cg23110422) and were associated with risk of UCC in the testing dataset (OR=1.37, 95% CI: 1.00-1.90) and (OR=1.42, 95% CI: 1.01-1.99), respectively. The association of MS10 with UCC risk in the WHI data (Model 2) was weaker (OR=1.09, 95% CI: 0.91-1.30).

**Table 4:** OR (per 1 SD increase), 95% CI and P value for the association between methylation-based predictors and risk of UCC.

| Predictor | Replication datasets |  |  |  |
| --- | --- | --- | --- | --- |
|  | MCCS (*N*=134 pairs) |  | WHI (*N*=440 pairs) |  |
|  | OR (95%CI) | P value | OR (95%CI) | P value |
| MS10 | 1.37 (1.00-1.90) | 0.05 | 1.09 (0.91-1.30) | 0.37 |
| MS66 | 1.35 (0.95-1.91) | 0.09 |  |  |
| MS11 | 1.42 (1.01-1.99) | 0.04 |  |  |
| MS2 | 1.05 (0.78-1.40) | 0.76 |  |  |
| MS18 |  |  | 1.09 (0.92-1.30) | 0.33 |

Note: The predictor was built by weighted average on methylation at selected CpGs: $MS = b_1CpG_1 + b_2CpG_2 + \dots + b_nCpG_n$ , where $CpG_i$ is M-value at this CpG site, $b_i$ use Lasso coefficients (for MS10, MS18) or log of OR from univariate analyses (for MS66, MS11, MS2). The association was estimated by conditional logistic regression Model 3 for MCCS data and Model 2 for WHI data, respectively.

Using all 404 case-control pairs of MCCS as the training set, as a sensitivity analysis, the logistic Lasso models selected 18 CpGs (MS18) from the 1,061 smoking-associated CpGs (Supplementary Table 7). MS18 and MS10 had eight overlapping CpGs (cg02743070, cg07058086, cg10399789, cg16622061, cg17924476, cg19089201, cg23110422 and cg24139443). We assessed the resulting predictor MS18 by examining its association with UCC risk in the WHI data, and the result was very similar as for MS10 (OR=1.09, 95% CI: 0.92-1.30) (Table 4). The fixed-effects meta-analysis for MS10 of the two replication sets in MCCS (*N*=134) and WHI (*N*=440) gave an estimated OR of 1.15, 95% CI: 0.98-1.34, P=0.08.

The ability of the methylation scores to predict risk of UCC with different models on the validation datasets is presented in Table 5. For the MCCS validation set, the predictions by Model C + MS10 and Model C + MS11 achieved the highest AUC estimate of 0.66, which was only slightly greater than the same model without methylation information (AUC=0.64, P=0.43 for MS10 and 0.39 for MS11). For the WHI testing set, the prediction by Model B + MS10 or MS18 achieved an AUC estimate of 0.68, which was of the same as Model B alone (P=0.11 or 0.22).

**Table 5:** AUC estimates and comparisons for predictions of UCC risk on the validation dataset sets using several models.

| MCCS (*N*=134 pairs) |  |  | WHI (*N*=440 pairs) |  |  |
| --- | --- | --- | --- | --- | --- |
|  | AUC | P value |  | AUC | P value |
| Model A | 0.61 | 0.18 (vs. Model C) | Model A | 0.58 | 0.0002 (vs. Model B) |
| Model B | 0.63 | 0.52 (vs. Model C) | Model B | 0.68 |  |
| Model C | 0.64 |  |  |  |  |
| Model A + MS10 | 0.63 | 0.27 (vs. Model A) | Model A + MS10 | 0.61 | 0.05 (vs. Model A) |
| Model A + MS11 | 0.64 | 0.19 (vs. Model A) | Model A + MS18 | 0.61 | 0.07 (vs. Model A) |
| Model B + MS10 | 0.65 | 0.36 (vs. Model B) | Model B + MS10 | 0.68 | 0.11 (vs. Model B) |
| Model B + MS11 | 0.65 | 0.30 (vs. Model B) | Model B + MS18 | 0.68 | 0.22 (vs. Model B) |
| Model C + MS10 | 0.66 | 0.44 (vs. Model C) |  |  |  |
| Model C + MS11 | 0.66 | 0.45 (vs. Model C) |  |  |  |

Note: The AUC was estimated based on unconditional logistic regression models. Model A used white blood cell composition as independent variables (for WHI, race/ethnicity was also used). Model B used white blood cell composition, smoking status and pack-years as independent variables (for WHI, race/ethnicity was also used). Model C used white blood cell composition, smoking status, pack-years and other covariates (age, sex, country of birth, sample type, alcohol, BMI, height, educational level, physical activity, socioeconomic status and diet quality) as independent variables. MS10, MS11 and MS18 were additional independent variables. P value was obtained by de Long test versus other models.

## Discussion

Most previous studies that investigated the association of smoking with development of urothelial cancer used self-reported smoking history. We included two self-reported variables smoking status and pack-years in our analyses. There are other aspects of smoking history, such as age at starting or passive smoking that are typically not or inaccurately captured by questionnaires. As DNA methylation in blood can capture lifetime exposure or different individual responses to smoking, we evaluated the association between smoking-associated methylation and risk of UCC. Although potential associations with UCC were identified at 206 (∼19%) and 93 (∼9%) smoking-based CpG sites in the MCCS in models without and with adjustment for self-reported smoking, respectively, and most associations were in the expected direction, these associations were overall quite weak. In the meta-analysis, DNA methylation at genes including *AHRR, GPR15, F2RL3, PRSS23* and *GFI1* (major smoking-related genes) was strongly (P<10^−7^) associated with UCC risk; however, the associations were substantially attenuated after adjusting for self-reported smoking history, likely because these self-reported variables might have captured almost full information of smoking exposure. Thus, these methylation markers added relatively little to the prediction of urothelial cancer risk beyond their association with self-reported smoking. A methylation score combining measures at ten smoking-associated CpG sites developed in the MCCS cohort showed some evidence of association with risk of UCC (OR per SD ∼ 1.4) independently of self-reported smoking in an independent dataset of MCCS participants (Table 4). Although these results suggest that the combination of smoking methylation markers may improve the prediction of urothelial cancer risk, limited evidence of replication was found in the WHI cohort (OR per SD ∼ 1.1).

The previous study by Jordahl et al. (24) using WHI data investigated three specific smoking-related probes (cg05575921 in the gene *AHRR*, cg03636183 in *F2RL3* and cg19859270 in *GPR15*) in relation to risk of UCC and showed that methylation alterations at cg05575921 and cg19859270 might mediate the effects of smoking on UCC. Our MCCS data also detected nominally significant associations with UCC risk at these CpGs (cg05575921: OR=0.78 [95% CI: 0.63-0.97], P=0.02 and cg19859270: OR=0.81 [95% CI: 0.68-0.97], P=0.02) in the adjusted model, which indicate they may add information about risk, in addition to the potential mediation of effect.

DNA methylation at *AHRR* cg05575921 was previously reported to be strongly associated with lung cancer risk (19, 34-36), e.g. OR=0.50 (95% CI: 0.43-0.59), P=4.3×10^−17^ in a pooled analysis of five case-control studies (19). Six CpGs in the *AHRR* gene also showed nominally significant association (P<0.05) with risk of UCC in our meta-analysis (Model 2): cg05575921 (OR=0.76, P=0.003), cg17924476 (OR=1.19, P=0.003), cg26529655 (OR=0.77, P=0.01), cg12806681 (OR=0.86, P=0.02), cg01899089 (OR=0.88, P=0.03) and cg03991871 (OR=0.88, P=0.04) (see Supplementary Table 5). Moreover, cg03636183 in the *F2RL3* gene, cg21566642 and cg05951221 in 2q37.1, and cg06126421 in 6p21.33 were also reported to be strongly associated (P=2×10^−15^) with lung cancer risk (19). Among them, three CpGs also showed nominally significant association with UCC risk in our meta-analyses (Model 2): cg21566642 (OR=0.82, P=0.009), cg05951221 (OR=0.86, P=0.04) and cg06126421 (OR=0.85, P=0.03) (see Supplementary Table 5). These associations appeared to be weaker than in the lung cancer studies, likely because smoking is not as strong a risk factor for urothelial cancer as it is for lung cancer. In a recent study (37), we showed that *GrimAge*, a composite biomarker based on several DNA methylation surrogates for plasma proteins and a methylation-based estimator of smoking pack-years (38), is substantially more strongly associated with lung cancer risk (OR per SD=2.03, 95% CI: 1.56-2.64) than with risk of UCC (OR=1.22, 95% CI: 0.98-1.52).

The samples used in the WHI cohort were all postmenopausal women and smoking accounts for approximately half of bladder cancer incidence among postmenopausal women (4, 23). Sex is associated with distinct DNA methylation patterns (39). However, we did not find that associations of DNA methylation smoking markers with UCC varied by sex in the MCCS data, nor did we find heterogeneity between MCCS and WHI results. In this study, we used two common methods to develop risk predictors: i) Lasso and ii) univariate analysis with weighted average based on individual CpG associations with UCC risk. For the latter, it is difficult to decide on an appropriate P-value cut-off and our results showed that the Lasso performed well in this setting. Although there was a reasonably large association of the Lasso predictor in the testing set (OR per 1 SD ∼ 1.4), this translated into only moderately improved risk prediction (Table 5).

There are several limitations in this study. First, even with pre-diagnostic blood samples, we cannot rule out the possibility that DNA methylation measures in blood reflected early cancer or development of other smoking-associated diseases. Second, the participants included in the MCCS testing set were an average eight years older than in the training set. As DNA methylation exhibits strong correlations with age, findings may have varied due to age differences between cohorts. We also noted that Model 1, which included only white blood cell composition variables, achieved an AUC of 0.53 for the training set but an AUC of 0.61 for the replication set (older MCCS participants). It may be that age, a strong cancer risk factor, is associated with changes in white blood cell composition over time (40) that are also associated with cancer risk (41, 42). Third, we considered the two MCCS datasets as independent because there was no participant overlap, and participants with follow-up blood samples were substantially older, however, the samples were drawn from the same cohort and might have shared environment; thus, the two datasets might not be completely independent, which may have an influence on results of validation and risk prediction. Fourth, the modest improvement of AUC may suggest that other factors, such as germline genetic variation, and incorporation of more environmental exposures, should be considered in the predictive models. Finally, compared with the MCCS cohort, the methylation measures in WHI were produced using different methods of sample collection and storage, DNA extraction, and DNA methylation processing, which may have influenced some findings, e.g. high heterogeneity for some CpGs across the two studies when performing meta-analysis.

In conclusion, our findings suggest that blood-based DNA methylation markers for smoking may be associated, albeit weakly, with risk of UCC independent of self-reported smoking history, and could provide some improvement to the prediction of urothelial cancer risk. The overall utility of our findings needs to be further assessed using additional external datasets.

## Supporting information

Supplementary Figure and Acknowledgements

## Data Availability

The data and code that support the findings of this study are available from the corresponding author upon reasonable request.

## Acknowledgements

We would like to acknowledge the Women’s Health Initiative (WHI) investigators. A short list of WHI investigators can be found in the supplementary data (Appendix 1).

## Authors’ contributions

C. Yu and P.A. Dugué conceived the study and designed the methodology. C. Yu, K.M. Jordahl, and P.A. Dugué conducted the data analysis. J.K. Bassett, J.E. Joo and E.M. Wong curated the data. J.E. Joo and E.M. Wong conducted laboratory work. R.L. Milne, P. Bhatti, M.C. Southey, and G.G. Giles contributed resources. P. Bhatti, M.C. Southey, G.G. Giles and P.A. Dugué obtained funding. P.A. Dugué supervised the study. C. Yu and P.A. Dugué drafted the manuscript. All authors contributed to the interpretation of the results and revision of the manuscript.

## References

1. Griffiths TL, Action on Bladder Cancer, Current perspectives in bladder cancer management. International Journal of Clinical Practice 2013; 67(5): 435–448.

2. Prasad SM, DeCastro GJ, Steinberg GD, Urothelial carcinoma of the bladder: definition, treatment and future efforts. Nature Reviews Urology 2011; 8(11): 631–642.

3. Bray F, Ferlay J, Soerjomataram I, Siegel RL, Torre LA, Jemal A, Global cancer statistics 2018: GLOBOCAN estimates of incidence and mortality worldwide for 36 cancers in 185 countries. CA: a cancer journal for clinicians 2018; 68(6): 394–424.

4. Freedman ND, Silverman DT, Hollenbeck AR, Schatzkin A, Abnet CC, Association between smoking and risk of bladder cancer among men and women. Jama 2011; 306(7): 737–745.

5. Burger M, Catto JW, Dalbagni G, Grossman HB, Herr H, Karakiewicz P, et al. Epidemiology and risk factors of urothelial bladder cancer. European urology 2013; 63(2): 234–241.

6. Crivelli JJ, Xylinas E, Kluth LA, Rieken M, Rink M, Shariat SF, Effect of smoking on outcomes of urothelial carcinoma: a systematic review of the literature. European urology 2014; 65(4): 742–754.

7. Hou L, Hong X, Dai M, Chen P, Zhao H, Wei Q, et al. Association of smoking status with prognosis in bladder cancer: a meta-analysis. Oncotarget 2017; 8(1): 1278.

8. Teleka S, Häggström C, Nagel G, Bjørge T, Manjer J, Ulmer H, et al. Risk of bladder cancer by disease severity in relation to metabolic factors and smoking: a prospective pooled cohort study of 800,000 men and women. International journal of cancer 2018; 143(12): 3071–3082.

9. Mori K, Mostafaei H, Abufaraj M, Yang L, Egawa S, Shariat SF, Smoking and bladder cancer: review of the recent literature. Current Opinion in Urology 2020; 30(5): 720–725.

10. van Osch FH, Jochems SH, van Schooten FJ, Bryan RT, Zeegers MP, Quantified relations between exposure to tobacco smoking and bladder cancer risk: a meta-analysis of 89 observational studies. International journal of epidemiology 2016; 45(3): 857–870.

11. Liber AC, Warner KE, Has underreporting of cigarette consumption changed over time? Estimates derived from US National Health Surveillance Systems between 1965 and 2015. American Journal of Epidemiology 2018; 187(1): 113–119.

12. Thomas CE, Wang R, Adams-Haduch J, Murphy SE, Ueland PM, Midttun Ø, et al. Urinary cotinine is as good a biomarker as serum cotinine for cigarette smoking exposure and lung cancer risk prediction. Cancer Epidemiology and Prevention Biomarkers 2020; 29(1): 127–132.

13. Benowitz NL, Biomarkers of environmental tobacco smoke exposure. Environmental health perspectives 1999; 107(suppl 2): 349–355.

14. Breitling LP, Yang R, Korn B, Burwinkel B, Brenner H, Tobacco-smoking-related differential DNA methylation: 27K discovery and replication. The American Journal of Human Genetics 2011; 88(4): 450–457.

15. Shenker NS, Ueland PM, Polidoro S, van Veldhoven K, Ricceri F, Brown R, et al. DNA methylation as a long-term biomarker of exposure to tobacco smoke. Epidemiology 2013; 24(5): 712–716.

16. Joehanes R, Just AC, Marioni RE, Pilling LC, Reynolds LM, Mandaviya PR, et al. Epigenetic signatures of cigarette smoking. Circulation: cardiovascular genetics 2016; 9(5): 436–447.

17. Zhang Y, Florath I, Saum KU, Brenner H, Self-reported smoking, serum cotinine, and blood DNA methylation. Environmental research 2016; 146: 395–403.

18. Besingi W, Johansson Å, Smoke-related DNA methylation changes in the etiology of human disease. Human molecular genetics 2014; 23(9): 2290–2297.

19. Baglietto L, Ponzi E, Haycock P, Hodge A, Bianca Assumma M, Jung CH, et al. DNA methylation changes measured in prelJdiagnostic peripheral blood samples are associated with smoking and lung cancer risk. International journal of cancer 2017; 140(1): 50–61.

20. Sabogal C, Su S, Tingen M, Kapuku G, Wang X, Cigarette smoking related DNA methylation in peripheral leukocytes and cardiovascular risk in young adults. International journal of cardiology 2020; 306: 203–205.

21. Marsit CJ, Koestler DC, Christensen BC, Karagas MR, Houseman EA, Kelsey KT, DNA methylation array analysis identifies profiles of blood-derived DNA methylation associated with bladder cancer. Journal of Clinical Oncology 2011; 29(9): 1133–1139.

22. Dugué PA, Brinkman MT, Milne RL, Wong EM, FitzGerald LM, Bassett JK, et al. Genome-wide measures of DNA methylation in peripheral blood and the risk of urothelial cell carcinoma: a prospective nested case–control study. British journal of cancer 2016; 115(6): 664–673.

23. Jordahl KM, Randolph TW, Song X, Sather CL, Tinker LF, Phipps AI, et al. Genome-Wide DNA Methylation in Prediagnostic Blood and Bladder Cancer Risk in the Women’s Health Initiative. Cancer Epidemiology and Prevention Biomarkers 2018; 27(6): 689–695.

24. Jordahl KM, Phipps AI, Randolph TW, Tindle HA, Liu S, Tinker LF, et al. Differential DNA methylation in blood as a mediator of the association between cigarette smoking and bladder cancer risk among postmenopausal women. Epigenetics 2019; 14(11): 1065–1073.

25. Milne RL, Fletcher AS, MacInnis RJ, Hodge AM, Hopkins AH, Bassett JK, et al. Cohort profile: the Melbourne collaborative cohort study (Health 2020). International journal of epidemiology 2017; 46(6): 1757–1757i.

26. Hays J, Hunt JR, Hubbell FA, Anderson GL, Limacher M, Allen C, et al. The Women’s Health Initiative recruitment methods and results. Annals of epidemiology 2003; 13(9): S18–S77.

27. Du P, Zhang X, Huang CC, Jafari N, Kibbe WA, Hou L, et al. Comparison of Beta-value and M-value methods for quantifying methylation levels by microarray analysis. BMC bioinformatics 2010; 11(1): 587.

28. Houseman EA, Accomando WP, Koestler DC, Christensen BC, Marsit CJ, Nelson HH, et al. DNA methylation arrays as surrogate measures of cell mixture distribution. BMC bioinformatics 2012: 13(1): 86.

29. Dugué PA, Jung CH, Joo JE, Wang X, Wong EM, Makalic E, et al. Smoking and blood DNA methylation: an epigenome-wide association study and assessment of reversibility. Epigenetics 2020: 15(4): 358–368.

30. Schwarzer G, meta: An R package for meta-analysis. R news 2007; 7(3): 40–45.

31. Friedman J, Hastie T, Tibshirani R, Regularization paths for generalized linear models via coordinate descent. Journal of statistical software 2010; 33(1): 1.

32. Robin X, Turck N, Hainard A, Tiberti N, Lisacek F, Sanchez JC, et al. pROC: an open-source package for R and S+ to analyze and compare ROC curves. BMC bioinformatics 2011; 12(1): 77.

33. DeLong ER, DeLong DM, Clarke-Pearson DL, Comparing the areas under two or more correlated receiver operating characteristic curves: a nonparametric approach. Biometrics 1988; 44: 837–845.

34. Fasanelli F, Baglietto L, Ponzi E, Guida F, Campanella G, Johansson M, et al. Hypomethylation of smoking-related genes is associated with future lung cancer in four prospective cohorts. Nature communications 2015; 6(1): 1–9.

35. Zhang Y, Breitling LP, Balavarca Y, Holleczek B, Schöttker B, Brenner H, Comparison and combination of blood DNA methylation at smoking-associated genes and at lung cancer-related genes in prediction of lung cancer mortality. International journal of cancer 2016; 139(11): 2482–2492.

36. Bojesen SE, Timpson N, Relton C, Smith GD, Nordestgaard BG, AHRR (cg05575921) hypomethylation marks smoking behaviour, morbidity and mortality. Thorax 2017; 72(7): 646–653.

37. Dugué PA, Bassett JK, Wong EM, Joo JE, Li S, Yu C, et al. Biological aging measures based on blood DNA methylation and risk of cancer: a prospective study. JNCI Cancer Spectrum 2021; 5(1): pkaa109.

38. Lu AT, Quach A, Wilson JG, Reiner AP, Aviv A, Raj K, et al. DNA methylation GrimAge strongly predicts lifespan and healthspan. Aging (Albany NY) 2019; 11(2): 303.

39. Boks MP, Derks EM, Weisenberger DJ, Strengman E, Janson E, Sommer IE, et al. The relationship of DNA methylation with age, gender and genotype in twins and healthy controls. PloS one 2009; 4(8): e6767.

40. Ruggiero C, Metter EJ, Cherubini A, Maggio M, Sen R, Najjar SS, et al. White blood cell count and mortality in the Baltimore Longitudinal Study of Aging. Journal of the American College of Cardiology 2007; 49(18): 1841–1850.

41. Erlinger TP, Muntner P, Helzlsouer KJ, WBC count and the risk of cancer mortality in a national sample of US adults: results from the Second National Health and Nutrition Examination Survey mortality study. Cancer Epidemiology and Prevention Biomarkers 2004; 13(6): 1052–1056.

42. Margolis KL, Rodabough RJ, Thomso, CA, Lopez AM, McTiernan A, Women’s Health Initiative Research Group, Prospective study of leukocyte count as a predictor of incident breast, colorectal, endometrial, and lung cancer and mortality in postmenopausal women. Archives of internal medicine 2007; 167(17): 1837–1844.

