## Supplementary Figure and Acknowledgements for "Smoking methylation marks for prediction of urothelial cancer risk"

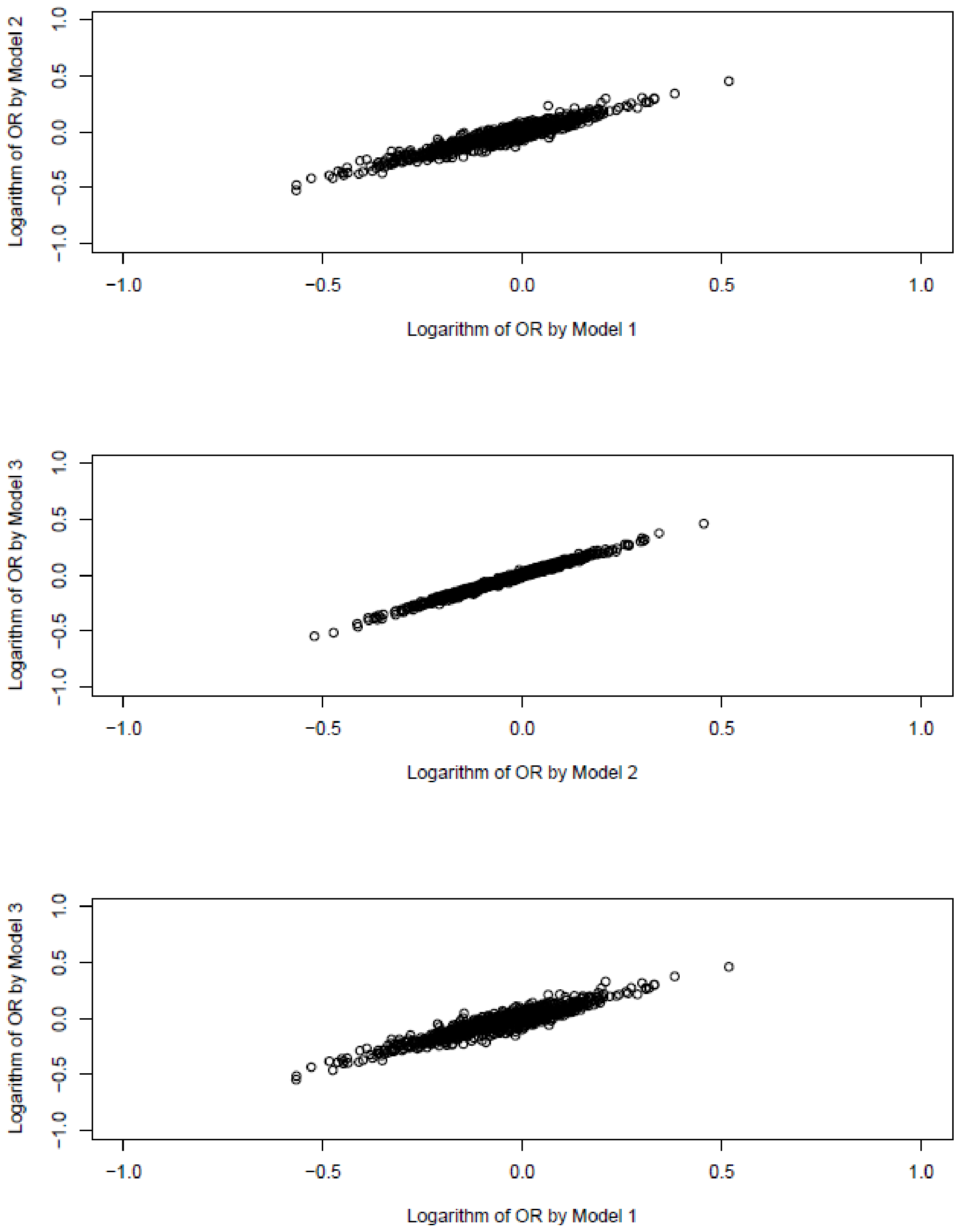


**Supplementary Figure 1**. Plots of regression coefficients in EWAS (404 matched case-control pairs in MCCS) at the 1,061 smoking-associated CpGs between Models 1-3.

**Appendix 1**. Short list of WHI investigators

**Program Office:** (National Heart, Lung, and Blood Institute, Bethesda, Maryland) Jacques Rossouw, Shari Ludlam, Joan McGowan, Leslie Ford, and Nancy Geller

**Clinical Coordinating Center:** (Fred Hutchinson Cancer Research Center, Seattle, WA) Garnet Anderson, Ross Prentice, Andrea LaCroix, and Charles Kooperberg

**Investigators and Academic Centers:** (Brigham and Women's Hospital, Harvard Medical School, Boston, MA) JoAnn E. Manson; (MedStar Health Research Institute/Howard University, Washington, DC) Barbara V. Howard; (Stanford Prevention Research Center, Stanford, CA) Marcia L. Stefanick; (The Ohio State University, Columbus, OH) Rebecca Jackson; (University of Arizona, Tucson/Phoenix, AZ) Cynthia A. Thomson; (University at Buffalo, Buffalo, NY) Jean Wactawski-Wende; (University of Florida, Gainesville/ Jacksonville, FL) Marian Limacher; (University of Iowa, Iowa City/Davenport, IA) Jennifer Robinson; (University of Pittsburgh, Pittsburgh, PA) Lewis Kuller; (Wake Forest University School of Medicine, Winston-Salem, NC) Sally Shumaker; (University of Nevada, Reno, NV) Robert Brunner

**Women's Health Initiative Memory Study**: (Wake Forest University School of Medicine, Winston-Salem, NC) Mark Espeland

For a list of all the investigators who have contributed to WHI science, please visit: *https://s3- us-west-2.amazonaws.com/www-whi-org/wp-content/uploads/WHI-Investigator-Long-List.pdf*
